# Surgical Critical Care Outcomes Before and After the Onset of the COVID-19 Pandemic: A Single-Center Retrospective Comparative Audit

**DOI:** 10.64898/2026.09.02.26361948

**Authors:** Syed Muhammad Hammad Ali, Umer Yasin, Ossama Ather, Asim Malik

## Abstract

This audit aimed to determine whether the pattern of presenting abdominal pathology, procedures performed, and clinical outcomes in our surgical intensive care unit (SICU) changed during the COVID-19 period. We reviewed a total of 356 patients who were admitted to the SICU with 1st March 2020 taken as the reference cutoff date. Two hundred and five (57.6%) cases occurred in the Pre-COVID interval and 151 (42.4%) in the Post-COVID interval. Patients admitted to SICU Post-COVID were significantly older than those admitted Pre-COVID (54.2 vs. 48.3 years, p = 0.005). Hospital and SICU length of stay were unchanged across the pandemic (p = 0.824 and 0.601 respectively). The procedure case-mix shifted significantly (p = 0.014): appendectomy, gastrectomy/sleeve gastrectomy and thoracic procedures fell, while exploratory laparotomy rose, consistent with reduced elective/minimally-invasive activity and a shift toward emergency open surgery. Appendiceal disease was recorded significantly less often as the presenting diagnosis Post-COVID (p = 0.016), and intestinal obstruction showed a non-significant upward trend (p = 0.071). SICU mortality and overall admission outcome were unchanged across the pandemic (p = 1.000 and 0.664).

## Background

The COVID-19 pandemic disrupted surgical services worldwide and reshaped how patients accessed emergency surgical care altogether. While global healthcare standards shifted to prioritizing attending to more emergent surgical cases, the fear of nosocomial infection, lockdown restrictions and disrupted primary-care pathways led many patients to delay presentation for common acute abdominal conditions. Multiple centers subsequently reported a higher proportion of complicated, perforated, or advanced disease at the time of surgery as demonstrated for acute appendicitis by Burgard et al.^1^ This pattern was also seen extending across the general emergency surgical case-mix, including cholecystitis and diverticulitis.^2^

However, most of the existing literature is drawn from single-condition case series (e.g. appendicitis alone, or mesenteric ischemia alone) rather than from a broader surgical intensive care population and evidence from our region/setting on this question is lacking. This audit was therefore undertaken to determine whether the pattern of presenting abdominal pathology, procedures performed, and clinical outcomes in our surgical ICU changed across the COVID-19 interval, and to explore further into whether any such shift is consistent with a pattern of change in disease severity and case-mix rather than an artefact of altered referral or admission practices.

## Methods

### Study design and population

This is a retrospective comparative audit of consecutive admissions to the surgical intensive care unit (SICU) at Fatima Memorial Hospital, drawn from prospectively recorded departmental audit records. Admissions were classified into two groups according to the timing of admission relative to the onset of the COVID-19 pandemic: “Pre-COVID” and “Post-COVID” based on the cut-off date of March 01, 2020. All surgical pre- or post-operative cases admitted six months prior to the cut-off date were categorized into the Pre-COVID group and all those admitted six months following the cut-off reference date were grouped into the Post-COVID category.

### Data source and variables

Data were extracted from a departmental audit spreadsheet recording, for each admission: timing of ICU admission: pre-operative (ICU admission prior to surgery, for optimization) or post-operative (ICU admission for post-operative recovery/monitoring), COVID interval, length of hospital stay (days), length of SICU stay (days), sex, age, free-text diagnosis, free-text procedure performed, admission outcome, and, where recorded, whether the patient required a second ICU admission within one month, with the diagnosis, procedure and outcome of that readmission.

### Data cleaning and standardization

Diagnosis and procedure fields were recorded as free text rather than according to a standardized classification (e.g. ICD-10); a formal ICD procedure code was present for only 39 of 356 records (11%) and was not used further. To enable quantitative comparison, diagnoses (213 distinct raw text entries) and procedures (189 distinct raw text entries) were independently grouped into a smaller number of clinically coherent categories using a reproducible, keyword-based rule set (e.g. all free-text variants of “cholelithiasis”, “choledocholithiasis”, “obstructive jaundice” and “cholecystitis” were grouped as “Biliary disease”; all variants of “CA”, “carcinoma”, “adenocarcinoma” and named sarcomas were grouped as “Malignancy (confirmed)”). This yielded 16 diagnosis categories and 27 procedure categories. This categorization was performed by the HA and OA and was cross checked by AM but it should be treated as a pragmatic approximation for hypothesis-generating analysis rather than a validated clinical classification for any future reference.

Eighteen of 356 records (5.1%) had a length-of-stay value meeting pre-specified implausibility criteria (negative value, value >200 days, or ICU stay exceeding hospital stay) and these individual values were treated as missing rather than the record being excluded entirely, since other fields for these records were unaffected.

### Statistical analysis

Descriptive statistics are presented as mean ± standard deviation or median with interquartile range (IQR) for continuous variables, depending on the distribution, and as frequency (%) for categorical variables. Continuous variables (age, hospital length of stay, SICU length of stay) were non-normally distributed and were compared between the Pre-COVID and Post-COVID groups using the Mann-Whitney U test. Categorical variables (sex, ICU admission timing, diagnosis category, procedure category, admission outcome) were compared using the chi-square test of independence; where more than 20% of expected cell counts were <5, categories were collapsed (rare categories combined into an “other” group) before testing, and Fisher’s exact test was used for all 2×2 comparisons and for post-hoc comparisons of individual diagnosis/procedure subcategories against all others. A two-sided p-value <0.05 was considered statistically significant. Because readmission status (“second ICU admission within one month”) was missing for a substantial and significantly unequal proportion of records in the two periods (see Results and Limitations), the readmission comparison is presented descriptively among records with the field recorded, with the differential missingness explicitly flagged as a source of potential bias rather than adjusted for. All analyses were performed in Python (SciPy v1.x).

### Missing Variables

Age was missing for 99/356 records (27.8%; 46 Pre-COVID, 53 Post-COVID); diagnosis for 32/356 (9.0%); procedure for 13/356 (3.7%). The “second ICU admission” field was missing for 203/356 records (57.0%) overall, and missingness differed significantly between periods (65.4% missing Pre-COVID vs. 45.7% missing Post-COVID, χ^2^ p<0.001), most likely reflecting a change in what was systematically recorded over the audit period rather than a true difference in follow-up completeness; readmission findings should be interpreted with this in mind. Diagnosis/procedure categorization was rule-based and not independently verified (see above).

## Results

### Baseline characteristics

A total of 356 patients were admitted to the SICU, 205 (57.6%) occurred in the Pre-COVID period and 151 (42.4%) in the Post-COVID period. Patients admitted in the Post-COVID period were significantly older than those admitted Pre-COVID (mean 54.2 vs. 48.3 years; Mann-Whitney U, p = 0.005). There was a non-significant trend toward a higher proportion of female patients Post-COVID (62.3% vs. 53.2%, χ^2^ p = 0.093). The proportion of admissions that were pre-operative vs. post-operative did not differ significantly between periods (χ^2^ p = 0.683). ***(Table 1)***

**Table 1.**
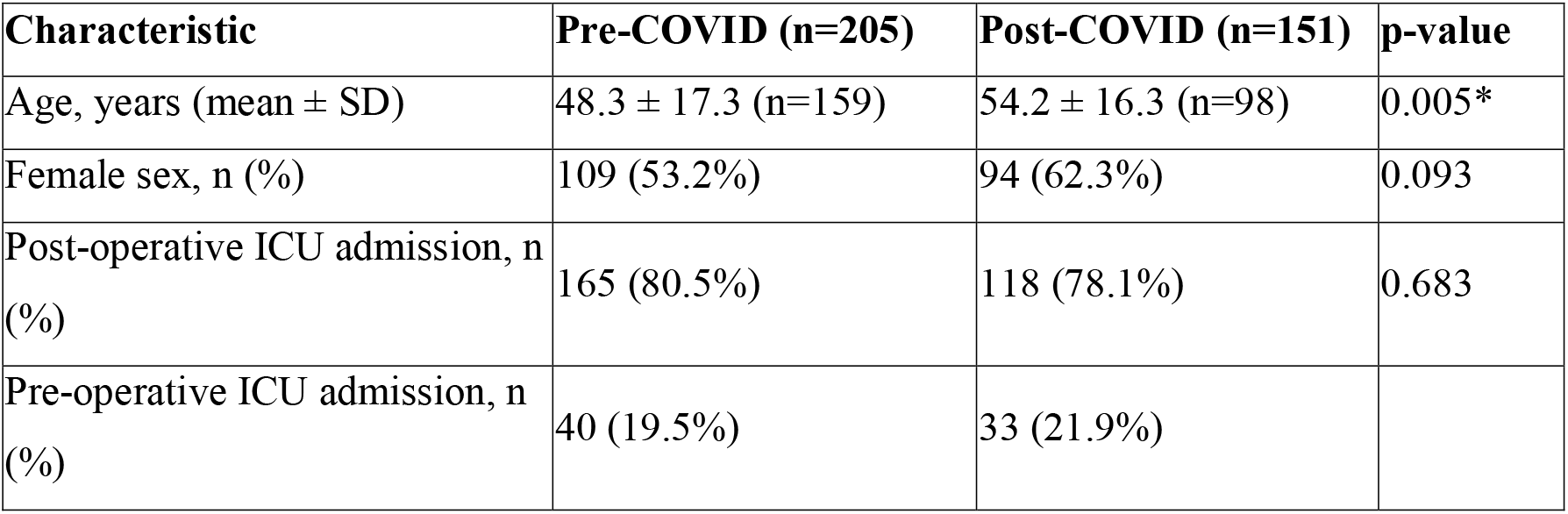
Baseline characteristics by COVID interval. *Statistically significant (p<0.05). SD = standard deviation.

| Characteristic | Pre-COVID (n=205) | Post-COVID (n=151) | p-value |
| --- | --- | --- | --- |
| Age, years (mean ± SD) | 48.3 ± 17.3 (n=159) | 54.2 ± 16.3 (n=98) | 0.005* |
| Female sex, n (%) | 109 (53.2%) | 94 (62.3%) | 0.093 |
| Post-operative ICU admission, n (%) | 165 (80.5%) | 118 (78.1%) | 0.683 |
| Pre-operative ICU admission, n (%) | 40 (19.5%) | 33 (21.9%) |  |

### Length of stay

Median hospital length of stay was 3.77 days (IQR 2.01–7.07) Pre-COVID and 4.00 days (IQR 1.95–6.68) Post-COVID; this difference was not statistically significant (Mann-Whitney U, p = 0.824). Median SICU length of stay was 1.95 days (IQR 0.97–4.15) Pre-COVID and 2.37 days (IQR 0.99–4.00) Post-COVID, not significantly different either (p = 0.601). Overall, the duration of critical care and hospital utilization per admission was essentially unchanged across the pandemic, despite the shift in age and case-mix described below. ***(Table 2)***

**Table 2.**
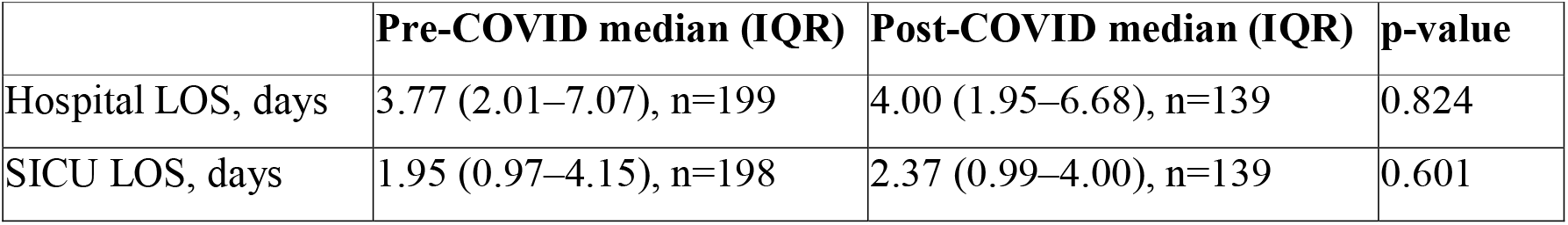
Length of stay by COVID interval. IQR = interquartile range; LOS = length of stay.

|  | Pre-COVID median (IQR) | Post-COVID median (IQR) | p-value |
| --- | --- | --- | --- |
| Hospital LOS, days | 3.77 (2.01–7.07), n=199 | 4.00 (1.95–6.68), n=139 | 0.824 |
| SICU LOS, days | 1.95 (0.97–4.15), n=198 | 2.37 (0.99–4.00), n=139 | 0.601 |

### Presenting diagnosis and procedure case-mix

The overall distribution of diagnosis categories did not reach statistical significance between periods (χ^2^ = 14.19, df = 8, p = 0.077, after collapsing categories with fewer than 15 total admissions). The distribution of procedure categories did differ significantly between periods (χ^2^ = 26.58, df = 13, p = 0.014, similarly collapsed), indicating a shift in the type of operative intervention performed across the pandemic. Post-hoc comparisons of individual categories against all others (Fisher’s exact test) identified the some statistically significant shifts. ***(Table 3)***

**Table 3.**
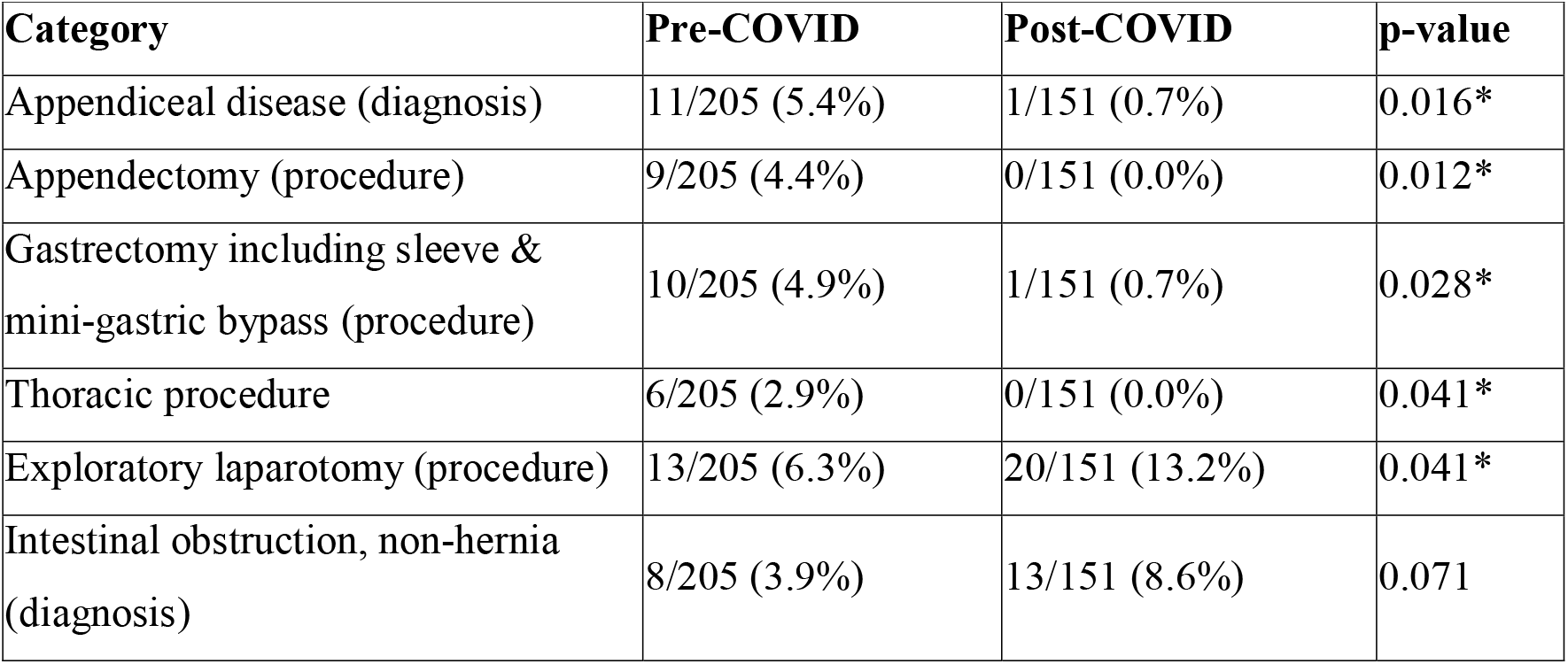
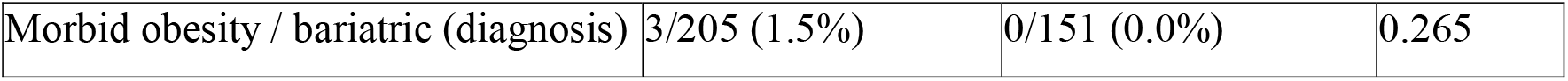
Individual diagnosis/procedure categories with a notable shift between periods (Fisher’s exact test, uncorrected for multiple comparisons). *Statistically significant (p<0.05).

In summary, elective and minimally invasive procedures like appendectomy, gastrectomy/sleeve gastrectomy, and thoracic procedures were significantly less frequent Post-COVID, while exploratory laparotomy (typically an open, urgent/emergency procedure) was significantly more frequent. Appendiceal disease was correspondingly less frequently recorded as the presenting diagnosis Post-COVID. Intestinal obstruction as a presenting diagnosis showed a non-significant trend toward being more frequent Post-COVID. This pattern is consistent with reduced provision of elective/minimally-invasive surgery and non-operative (conservative) management of some acute surgical presentations (e.g. appendicitis) during the pandemic period, with a compensatory shift toward more open, urgent surgery for those who did proceed to theatre, a pattern that has been reported in surgical practice internationally during COVID-19 and would be worth discussing explicitly in relation to your own department’s pandemic-era protocols.^3^

### Admission outcome and mortality

Overall admission outcome (discharged home, discharged to inpatient ward/High Dependency Unit (HDU), expired, transferred, left against medical advice, other) did not differ significantly between periods (χ^2^ = 3.23, df = 5, p = 0.664). SICU mortality was similar between periods: 21/205 (10.2%) Pre-COVID vs. 16/151 (10.6%) Post-COVID (Fisher’s exact, OR 0.96, p = 1.000). ***(Table 4)***

**Table 4.** Admission outcome by COVID interval (χ^2^ p = 0.664 across all categories; mortality alone Fisher’s p = 1.000). HDU: High Dependency Unit, LAMA: Left Against Medical Advice

| Outcome | Pre-COVID, n (%) | Post-COVID, n (%) |
| --- | --- | --- |
| Discharged to ward/HDU | 135 (65.9%) | 92 (60.9%) |
| Discharged home | 47 (22.9%) | 37 (24.5%) |
| Expired | 21 (10.2%) | 16 (10.6%) |
| LAMA / transferred / other | 2 (1.0%) | 6 (4.0%) |

### Readmission

Among the subset of records with readmission status recorded (71/205 Pre-COVID, 82/151 Post-COVID), a second ICU admission within one month occurred in 5/71 (7.0%) Pre-COVID vs. 5/82 (6.1%) Post-COVID (Fisher’s exact, OR 1.17, p = 1.000), no evidence of a difference. However, this field was missing far more often Pre-COVID (65.4%) than Post-COVID (45.7%), a highly significant difference in recording completeness (χ^2^ p<0.001) rather than a clinical finding.

## Summary of key findings

1. Patients admitted to SICU Post-COVID were significantly older than those admitted Pre-COVID (54.2 vs. 48.3 years, p = 0.005).
2. Hospital and SICU length of stay were unchanged across the pandemic (p = 0.824 and 0.601 respectively).
3. The procedure case-mix shifted significantly (p = 0.014): appendectomy, gastrectomy/sleeve gastrectomy and thoracic procedures fell, while exploratory laparotomy rose, consistent with reduced elective/minimally-invasive activity and a shift toward emergency open surgery.
4. Appendiceal disease was recorded significantly less often as the presenting diagnosis Post-COVID (p = 0.016), and intestinal obstruction showed a non-significant upward trend (p = 0.071).
5. SICU mortality and overall admission outcome were unchanged across the pandemic (p = 1.000 and 0.664).

## Data Availability

All data produced in the present study are available upon reasonable request to the authors

## References

1. Burgard M, Cherbanyk F, Nassiopoulos K, Malekzadeh S, Pugin F, Egger B. An effect of the COVID-19 pandemic: significantly more complicated appendicitis due to delayed presentation of patients! PLoS One. 2021;16(5):e0249171. 10.1371/journal.pone.0249171

2. Vissio E, Falco EC, Scozzari G, Scarmozzino A, Trinh DAA, Morino M, Papotti M, Bertero L, Cassoni P. The adverse impact of the COVID-19 pandemic on abdominal emergencies: a retrospective clinico-pathological analysis. J Clin Med. 2021;10(22):5254. 10.3390/jcm10225254

3. Kronberga M, Saha A. SP11.3 Impact of Covid on emergency laparotomy activity. Br J Surg. 2022 Aug 9;109 (Suppl 5):znac247.122. 10.1093/bjs/znac247.122

